# Neuropsychiatric copy number variants exert shared effects on human brain structure

**DOI:** 10.1101/2020.04.15.20056531

**Authors:** Claudia Modenato, Kuldeep Kumar, Clara Moreau, Sandra Martin-Brevet, Guillaume Huguet, Catherine Schramm, Jean-Louis Martineau, Charles-Olivier Martin, Nadine Younis, Petra Tamer, Elise Douard, Fanny Thébault-Dagher, V. Côté, A.R. Charlebois, F. Deguire, Anne M. Maillard, Borja Rodriguez-Herreros, Aurèlie Pain, Sonia Richetin, 16p11.2 European Consortium, Simons Variation in Individuals Project (VIP) Consortium, Leila Kushan, Lester Melie-Garcia, Ana I. Silva, Marianne B.M. van den Bree, David E.J. Linden, Michael J. Owen, Jeremy Hall, Sarah Lippé, Mallar Chakravarty, Danilo Bzdok, Carrie E. Bearden, Bogdan Draganski, Sébastien Jacquemont

## Abstract

**Background:** Copy Number Variants (CNVs) associated with autism and schizophrenia have large effects on brain anatomy. Yet, neuroimaging studies have been conducted one mutation at a time. We hypothesize that neuropsychiatric CNVs may exert general effects on brain morphometry because they confer risk for overlapping psychiatric conditions.

**Methods:** We analyzed T1-weighted MRIs and characterized shared patterns on brain anatomy across 8 neuropsychiatric CNVs. Clinically ascertained samples included 1q21.1 (n=48), 16p11.2 (n=156), or 22q11.2 (n=96) and 331 non-carriers. Non-clinically ascertained samples from the UK Biobank included 1q21.1 (n=19), 16p11.2 (n=8), 22q11.2 (n=9), 15q11.2 (n=148) and 965 non-carriers. Canonical correlation analysis (CCA) and univariate models were used to interrogate brain morphometry changes across 8 CNVs.

**Results:** Eight CNVs affect regional brain volumes along two main gene-morphometry dimensions identified by CCA. While fronto-temporal regions contributed to dimension 1, dimension 2 was driven by subcortical, parietal and occipital regions. Consistently, voxel-wise whole-brain analyses identified the same regions involved in patterns of alteration present across the 4 deletions and duplications. These neuroanatomical patterns are similar to those observed in cross-psychiatric disorder meta-analyses. Deletions and duplications at all 4 loci show mirror effects at either the global and/or the regional level.

**Conclusion:** Neuropsychiatric CNVs share neuroanatomical signatures characterized by a parsimonious set of brain dimensions. The latter may underlie the risk conferred by CNVs for a similar spectrum of neuropsychiatric conditions.

## Introduction

Genomic copy number variants (CNVs) are deletions or duplications of DNA segments of more than 1000 base pairs. Rare CNVs with large effects have been associated with a range of neurodevelopmental and psychiatric conditions. Sixteen recurrent CNVs have been associated with autism spectrum disorder (ASD) ^1^ and eight with schizophrenia (SZ) ^2^. Among those, CNVs at the 22q11.2 (BPA-D), 16p11.2 (BP4-5), 1q21.1 (Class I & II) and 15q11.2 (BP1-2) genomic loci are the most frequently identified risk-factors for neuropsychiatric disorders identified in the clinic ^3,4^. They affect the dosage of 60, 29, 12 and 4 genes, respectively ^5–7^. The largest increases in risk for SZ have been documented for the 22q11.2 deletion (30 to 40-fold) followed by the 16p11.2 duplication (10-fold), the 1q21.1 deletion and the 15q11.2 deletion (1.5-fold) ^2^. ASD risk is highest for 16p11.2 deletions and duplications (10-fold) followed by 1q21.1 duplications and 22q11.2 duplications (3 to 4-fold) ^1,2,8–11^. These variants are therefore opportunities to investigate the brain phenotypes associated with high-risk for mental illness.

Neuroimaging studies have only been performed for a few CNVs. They have shown robust effects on total and regional brain volumes e.g. 22q11.2 ^10,12^, 16p11.2 BP4-5 ^13–15^, and 15q11.2 ^16–19^. Deletions and duplications at the 16p11.2, and to a lesser extent 22q11.2, loci affect global and regional brain volumes in opposite directions. This is also observed for cortical thickness (CT) and white matter (DTI) in deletions and duplications of the 15q11.2 BP1-BP2 locus ^17,19^. Moreover, neuroanatomical alterations associated with 16p11.2 and 22q11.2 show overlap with those observed in idiopathic ASD and SZ ^10,13–15,17.^

However, CNV studies have been conducted one mutation at a time, which hinders our understanding of any potential general mechanism linking CNVs to effects on brain architecture. Indeed, several lines of evidence suggest shared mechanism across CNVs: 1) Neuropsychiatric CNVs have been associated with a similar range of adverse effects on childhood neurodevelopment with only subtle quantitative and qualitative differences^20^, 2) CNVs affecting coding genes decrease IQ and increase risk for ASD across a large proportion of the genome ^21,22^, 3) Schizophrenia is associated with an increase in overall CNV burden ^2^.

The large number of genes encompassed in CNVs has also limited the study of mechanisms associating CNVs to brain morphometry. Studies of the 16p11.2 locus have suggested that TAOK2 and KCTD13 within the 16p11.2 locus are implicated in embryonic neurogenesis and are candidate genes for differences in total brain volume in animal models ^6,23^. Similarly, DGCR8 and TBX1 within the 22q11.2 locus were found to be involved in neurogenesis ^24,25^. The effects of these genes on regional volumes have not yet been studied in humans or animal models.

The body of literature on CNVs raises several questions: What is the relationship between global and regional alterations associated with CNVs? Does gene dosage at distinct genomic loci converge on a common set of brain alterations, or do genes lead to specific effects? More broadly, the field is lacking a conceptual framework to identify general principles linking CNVs to effects on brain architecture and risk for psychiatric conditions.

In this study, we aimed to characterize shared and specific neuroanatomical variations across multiple CNVs by capitalizing on multivariate and univariate methods.

We analyzed T1 weighted MRIs of the largest multi-site dataset of CNV carriers (n=484, of which 87 have not yet been published) and controls (n=1296). Voxel and surface-based methods were performed in parallel. First, a multi-view pattern-learning algorithm (Canonical Correlation Analysis, CCA) was used to identify latent brain morphometry dimensions explaining the effects of CNVs across genomic loci. Second, we investigated brain alterations shared across deletions and duplications using univariate linear models.

## Methods

### Clinically ascertained CNV carriers

Individuals carrying 1q21.1 (Class I & II), 22q11.2 (BPA-D) or 16p11.2 (BP4-5) CNVs, were assessed as either probands referred for genetic testing, or as relatives. Controls were either non-carriers within the same families or individuals from the general population. We pooled data from 5 cohorts: Cardiff University (UK), 16p11.2 European Consortium (Lausanne, Switzerland), University of Montreal (Canada), UCLA (Los Angeles, USA) and the Variation in individuals Project (SVIP, USA) detailed in the supplementary files.

### CNVs assessed in non-clinical populations

Genetic and neuroimaging data from non-clinical population were obtained from the UK Biobank dataset. PennCNV and QuantiSNP were used, using standard quality control metrics, to identify CNVs ^21,22^. Deletions and duplications with neuroimaging data included in the study were selected on the following breakpoints: 16p11.2 (BP4-5, 29.6-30.2MB), 1q21.1 (Class I, 146.4-147.5MB & II, 145.3-147.5MB), 22q11.2 (BPA-D, 18.8-21.7MB) and 15q11.2 (BP1-2, 22.8-23.0MB), together with control individuals not carrying any CNVs at these loci (Table 1 and eTable 1). Signed consents were obtained from all participants or legal representatives prior to the investigation.

**Table 1.** Demographics. CNV: Copy Number Variant, SD: Standard deviation, TIV: total intracranial volume, FSIQ: Full scale IQ, UKB FI: UK Biobank fluid intelligence, ASD: Autism Spectrum Disorders, SCZ: Schizophrenia (including * ICD10 code F25.9 Schizoaffective disorder, unspecified). CNV carriers and controls from the clinically ascertained group come from 5 different cohorts (eTable 1), while non-clinically ascertained participants were identified in the UK Biobank. UK Biobank fluid intelligence scores (UKB field:20016) were adjusted for age, age^2^, sex, site and then z-scored. 16p11.2 and 22q11.2 from the UKBB were not included in the VBM and SBM due to small sample size.

| CLINICAL ASCERTAINMENT |  |  |  |  |  |  |  |  |
| --- | --- | --- | --- | --- | --- | --- | --- | --- |
| CNV loci | Copy number | Age mean (SD) | Male/ Female | TIV mean (SD) | FSIQ mean (SD) | ASD | SCZ | Other diagnosis |
| 1q21.1 | Deletions N=29 | 29(18) | 11/18 | 1.22(0.14) | 90.85 (21.75) N=26 | 1 | - | 7 |
|  | Duplication N=19 | 34(17) | 10/9 | 1.57(0.11) | 95.56 (23.19) N=18 | 1 | - | 4 |
| 16p11.2 | Deletions N=83 | 17(12) | 47/36 | 1.54(0.17) | 82.17 (14.99) N=64 | 13 | - | 36 |
|  | Duplication N=73 | 31(14.9) | 41/32 | 1.33(0.17) | 85.47 (19.48) N=63 | 10 | 1 | 19 |
| 22q11.2 | Deletions N=74 | 16(8.6) | 35/39 | 1.30(0.15) | 77.42 (13.51) N=48 | 9 | 2 | 32 |
|  | Duplication N=22 | 20(14.2) | 15/7 | 1.47(0.16) | 97.83 (20.34) N=12 | 2 | - | 8 |
| Controls N=331 |  | 26(14.6) | 189/142 | 1.46(0.15) | 106.73 (15.03) N=224 | 1 | - | 23 |
| NON-CLINICAL ASCERTAINMENT |  |  |  |  |  |  |  |  |
| CNV loci | Copy number | Age mean (SD) | Male/ Female | TIV mean (SD) | UKB FI mean (SD) | ASD | SCZ | Other diagnosis |
| 1q21.1 | Deletions N=10 | 59.1(6.7) | 6/4 | 1.35(0.12) | -0.8 (0.5) N=9 | - | 1* | 3 |
|  | Duplication N=9 | 60.6(7) | 2/7 | 1.55(0.14) | 0.2 (1.3) N=9 | - | - | - |
| 15q11.2 | Deletions N=72 | 63.4(7.6) | 31/41 | 1.54(0.15) | -0.3 (0.9) N=63 | - | - | 2 |
|  | Duplication N=76 | 62.9 (7.3) | 36/40 | 1.49(0.15) | 0 (1.1) N=71 | - | - | 6 |
| 16p11.2 | Deletion N=4 | 65.6 (3.2) | 3/1 | 1.56(0.13) | 0.8 (0.5) N=2 | - | - | - |
|  | Duplication N=4 | 69.3 (2.1) | 1/3 | 1.29(0.11) | -1.6 (0.2) N=4 | - | - | - |
| 22q11.2 | Deletion N=1 | 69.8(-) | 1/- | 1.44(-) | - | - | - | - |
|  | Duplication N=8 | 62(9.5) | 4/4 | 1.55(0.17) | -0.2 (1.1) N=8 | - | - | 1 |
| Controls N=965 |  | 62.1(7.4) | 358/607 | 1.51(0.14) | 0 (1) N=866 | - | 2* | 65 |

### MRI data

Data sample included T1-weighted (T1w) images at 0.8 −1 mm isotropic resolution across all sites. The data distribution overview, as well as the population description is available in Table 1 and eTable 1. MRI protocol information is available in supplementary information.

### Data quality check

All data used in the analysis were quality checked by the same researcher (CM). A total of 107 structural brain scans were excluded from further analysis based on standardized visual inspection that identified significant artefacts compromising the accurate tissue classification and boundary detection (details in supplementary materials and methods).

### MRI data processing, Voxel-Based Morphometry

Data were preprocessed and analysed with SPM12 (http://www.fil.ion.ucl.ac.uk/spm/software/spm12/) running under MATLAB R2018b (https://www.mathworks.com/products/new_products/release2018b.html). Preprocessing steps are described in detail in supplementary eMethod 1.

### MRI data processing, Surface-Based Morphometry

Brain scans were processed using FreeSurfer 5.3.0 (http://surfer.nmr.mgh.harvard.edu ^29^). Quality control involved visual inspection of each cortical surface reconstruction output (KK) and the standardized ENIGMA quality control procedures (http://enigma.ini.usc.edu/protocols/imaging-protocols/). Detailed procedure is provided in supplementary eMethod 2.

### Statistical analysis for global brain measures - Total Intracranial Volume (TIV), total Gray Matter volume (GM), total Surface Area (SA) and mean Cortical Thickness (CT)

Global brain aggregate measures were adjusted for age, age^2^, and sex as fixed effects and scanning site as a random factor. Global measure z-scores for each CNV for clinically and non-clinically ascertained CNVs were calculated using 331 and 965 controls, respectively. Within the two cohort ascertainments, we used ANOVA design to compare group means and p-value correction for multiple comparisons was performed with the Tukey Honest Significant Differences test. Wilcoxon Rank Sum and Signed Rank Tests were used to compare distributions between deletions and duplications of each CNV. All statistical analyses were performed in R, version 3.4.4 (https://www.r-project.org/) or in MatlabR2018b.

### Multi-view pattern-learning analysis

We re-purposed canonical correlation analysis (CCA) to interrogate the shared and distinct impact on brain morphometry (130 grey matter regions) across CNVs ^31,32^. This principled multivariate approach allowed investigating the underlying relationship between two sets of variables, and has been widely used in neuroimaging studies ^31,32^. In our study, CCA identified modes of coherent co-variation that jointly characterize how CNVs and patterns of regional volumes systematically co-occur across subjects. We refer to these modes of co-variation as ‘CCA dimensions’ or ‘gene-morphology dimensions’. Details are provided in supplementary eMethod 3.

### Voxel-based measures and statistical analyses

To complement and illustrate the CCA analysis, we performed a whole-brain voxel-based approach. Analyses tested voxel-wise differences in volume using a mass-univariate analysis framework implemented in SPM. Details are available in supplementary eMethod 4. Cohen’s d (i.e. effect-size) ^35^ maps were obtained by converting SPM T-maps using the CAT12 toolbox for SPM (http://www.neuro.uni-jena.de/cat/).

### Surface-based measures and statistical analyses

In parallel to VBM, we used surface-based GLM-based analysis to test differences in CT and SA (SurfStat toolbox ^36^). Each GLM used the surface feature as the dependent variable, the groups - as independent variable, which were adjusted for age, age^2^, sex, site, and Total-SA/Mean-CT. Post-hoc contrasts compared each CNV group against controls, including estimation of Cohen’s d effect size estimates from t-values ^35^. False Discovery Rate (FDR), with p-value at 0.05 ^12–15^ was applied to control for false positive errors due to multiple comparisons.

### Ranked Cohen’s d maps compared across CNVs

To adjust for the unequal power to detect change across different CNV groups in the univariate analyses, which have different sample and effect sizes, we ranked Cohen’s d distributions of all voxels (/vertices) from the estimated un-thresholded statistical maps. We then tested for spatial overlap between maps across CNVs after thresholding the tails of the distribution at: i) 5th & 95^th^ quantiles, and ii) 15th & 85^th^ quantiles. The dice-index was calculated using publicly available Matlab scripts and functions (https://github.com/rordenlab/spmScripts).

### Comparison across VBM, SA, and CT

To compare spatial pattern of voxel- and surface-based results, we projected VBM results on fsaverage, using Freesurfer’s vol2surf function.

### Null hypotheses using spin permutations and label shuffling

We used spin permutation and label shuffling to calculate empirical p-values for 1) the deletion and duplication convergence pattern and 2) the correlation/dice-index between two maps.

### Spin permutation testing

The spin permutation test provides a null hypothesis quantifying the probability of observing by chance a dice index or correlation value, while controlling for spatial auto-correlations inherent in neuroimaging data. This method has been established previously in ^37,38^, and detailed in supplementary eMethod 5.

### Label shuffling

We additionally tested the overlap significance by performing permutation of control and CNV labels, generating empirical null distributions and calculating dice index distribution with regard to the convergence pattern. Details are available in supplementary eMethod 6.

### Overlap with cross-psychiatric disorders map

Dice-index was computed to estimate the overlap between deletion and duplication convergence maps and the statistical maps obtained from a large cross-disorder neuroimaging meta-analysis (http://anima.fz-juelich.de) ^39^.

## Results

### 1. Effects on global brain morphometry

Deletions and duplications of each genomic loci (except 15q11.2) showed opposing effects on total intracranial volume (TIV), total grey matter volume (GM) and total surface area (SA) (Figure 1a-c). For mean cortical thickness (CT), 22q11.2 and 15q11.2 deletions and duplications showed opposing effects (Figure 1d). The directionality of global effects differed across loci: They were positively correlated with gene dosage at the 1q21.1, 22q11.2 and 15q11.2 loci (i.e., duplications associated with greater TIV, SA or CT), and negatively correlated with gene dosage at the 16p11.2 locus (Figure 1a-c.). Similar mirror effects on TIV, GM volume and total SA (Figure 1a-c.) in 1q21.1 CNV carriers recruited in the clinic and identified in the UK Biobank, indicated that global effects were not influenced by ascertainment.

**Figure 1:**
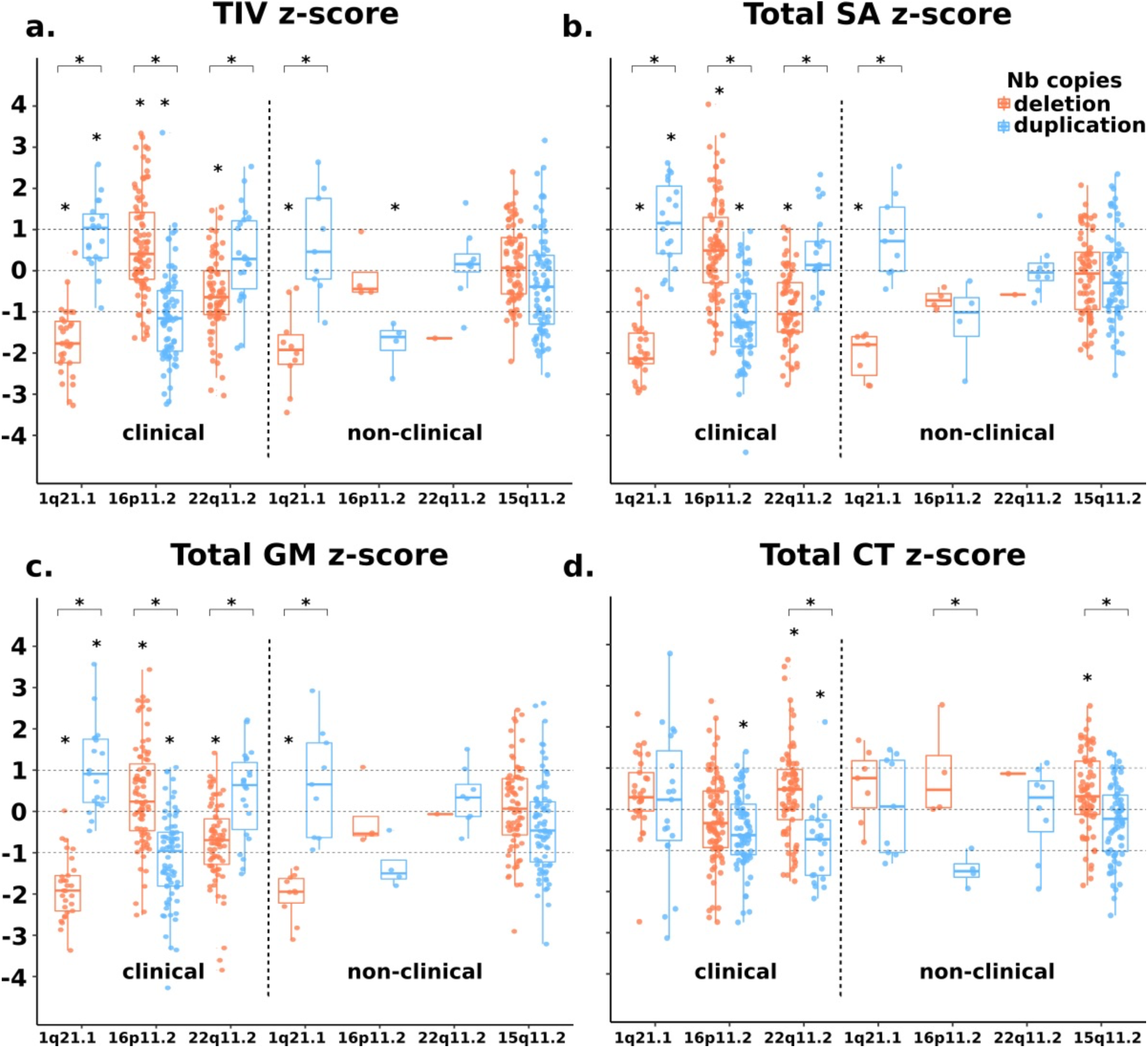
The effect of 1q21.1, 16p11.2, 22q11.2 and 15q11.2 on global brain measures. Total intracranial (a), total surface area (b), total grey matter volume (c) and mean cortical thickness (d) for clinically and non-clinically ascertained CNVs. Z-scores for clinically and non-clinically ascertained CNVs were calculated using 331 and 965 controls respectively, adjusting for age, age^2^, sex and site as a random factor. Y axis values are z scores. X axis are CNV groups. Significant difference between CNV group and corresponding control group is indicated with a star. Horizontal bars with stars show significant differences between deletions and duplications within the same locus.

### 2. Brain morphometry dimensions across 8 CNVs

We used canonical correlation analysis (CCA) to interrogate morphometry changes of 130 regional grey matter volumes across 4 genomic loci. Deletions and duplications were coded as opposing gene dosage at each locus. CCA was calculated using 484 CNV carriers without controls and identified 3 ‘gene-morphology dimensions’ (r=0.84, 0.79, 0.73; statistically significant at p-value<0.01). Top ranking brain regions contributing to the most dominant dimensions of morphological variation included transverse temporal gyrus, planum temporale, parietal operculum/calcarine cortex, supplementary motor cortex, posterior and anterior insula, middle cingulate and temporal gyrus. Those contributing most to dimension 2 of gene-morphology co-variation included the accumbens, subcallosal area, cuneus, precuneus, brain stem, superior frontal gyrus, cerebellum, anterior insula and lingual gyrus (Figure 2b-c, eTable 5). 16p11.2 and 22q11.2 preferentially contributed to dimension 1 and 2 respectively, and 1q21.1 loaded similarly on both dimensions. 15q11.2 CNVs showed the smallest loadings on both dimensions (Figure 2d).

**Figure 2:**
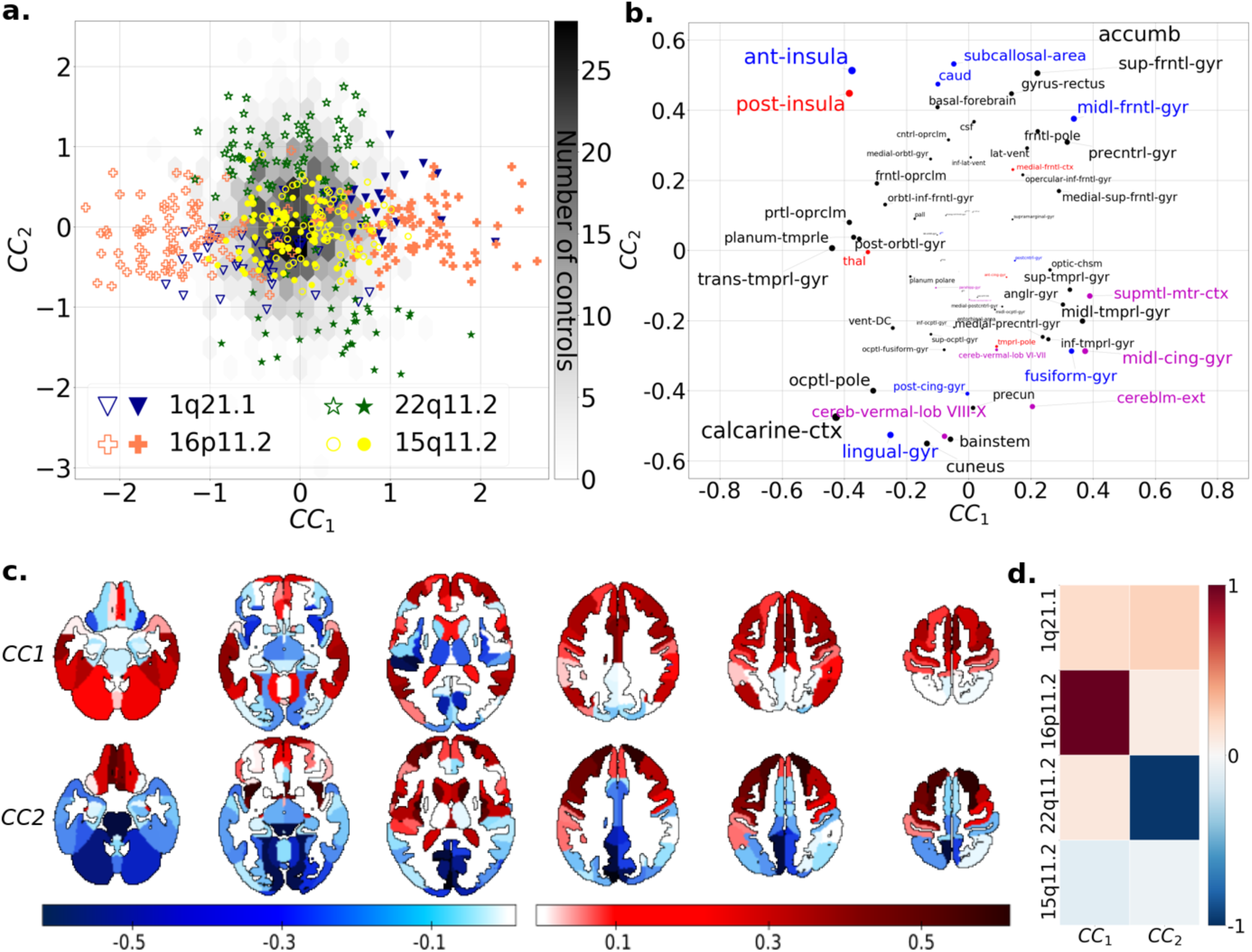
Co-analysis of shared brain changes due to 8 CNVs. (a) Scatterplot showing the position of each of the 484 carriers of 8 different CNVs along 2 dominant brain-gene Canonical Correlation (CC) dimensions established using 130 neuroanatomical GM regions of CNV carriers. GM region volumes were obtained using neuromorphometric and were adjusted for total grey matter, age, age^2^ sex and site. The empty and full symbols represent deletions and duplication respectively. The grey hexagonal bin plot represents the frequency of controls (n=1296). Controls were not used to calculate the CCA and were projected post hoc on the 2 dimensions using CCA prediction. X and Y axis values: z-scores of regional volumes. (b) Loading of Neuromorphometric Regions of Interests (ROIs) on the 2 CC dimensions. X axis and Y axis values: normalized values from (−1 to +1) of dimensions 1 and 2 respectively. The font size is correlated to the region’s contribution to dimensions. ROI names are color coded as being part of the deletion (red), duplication (blue) and both deletion and duplication (magenta) convergence patterns. (c) CCA dimension 1 and 2 regional relevances projected on axial brain slices. The darker the red or blue color, the stronger the positive or negative association with the CCA dimensions. (d) Loading of the first and second CCA dimension on 4 CNV genomic loci. Values are CCA loading magnitudes and represent the contribution of a CNV loci to the canonical dimension.

Projecting each CNV carrier on the canonical dimensions demonstrated contrasting loadings between deletions and duplications, and illustrated the ranking of effect-sizes for each genomic locus (From largest to smallest: 22q11.2, 16p11.2, 1q21.1 and 15q11.2, Figure 2a, supplementary eTable 2-4). While the two gene-morphology dimensions allowed for discrimination between genomic loci and between most CNV carriers and controls, they also pointed to brain regions contributing to variation shared across loci (Figure 2b).

### 3. Deletions at 4 genomic loci converge on shared neuroanatomical alterations

To further dissect the “gene-morphology dimensions” identified above, we performed univariate whole-brain VBM analyses contrasting each deletion and duplication group with controls. We first only considered clinically ascertained individuals to account for selection bias. Cohen’s d maps from the 22q11.2 and 16p11.2 CNVs were consistent with previous studies (Figure 3) ^10,15^. Findings for all 3 deletions including 1q21.1 are detailed in Figure 3 and supplementary eTable 2.

**Figure 3:**
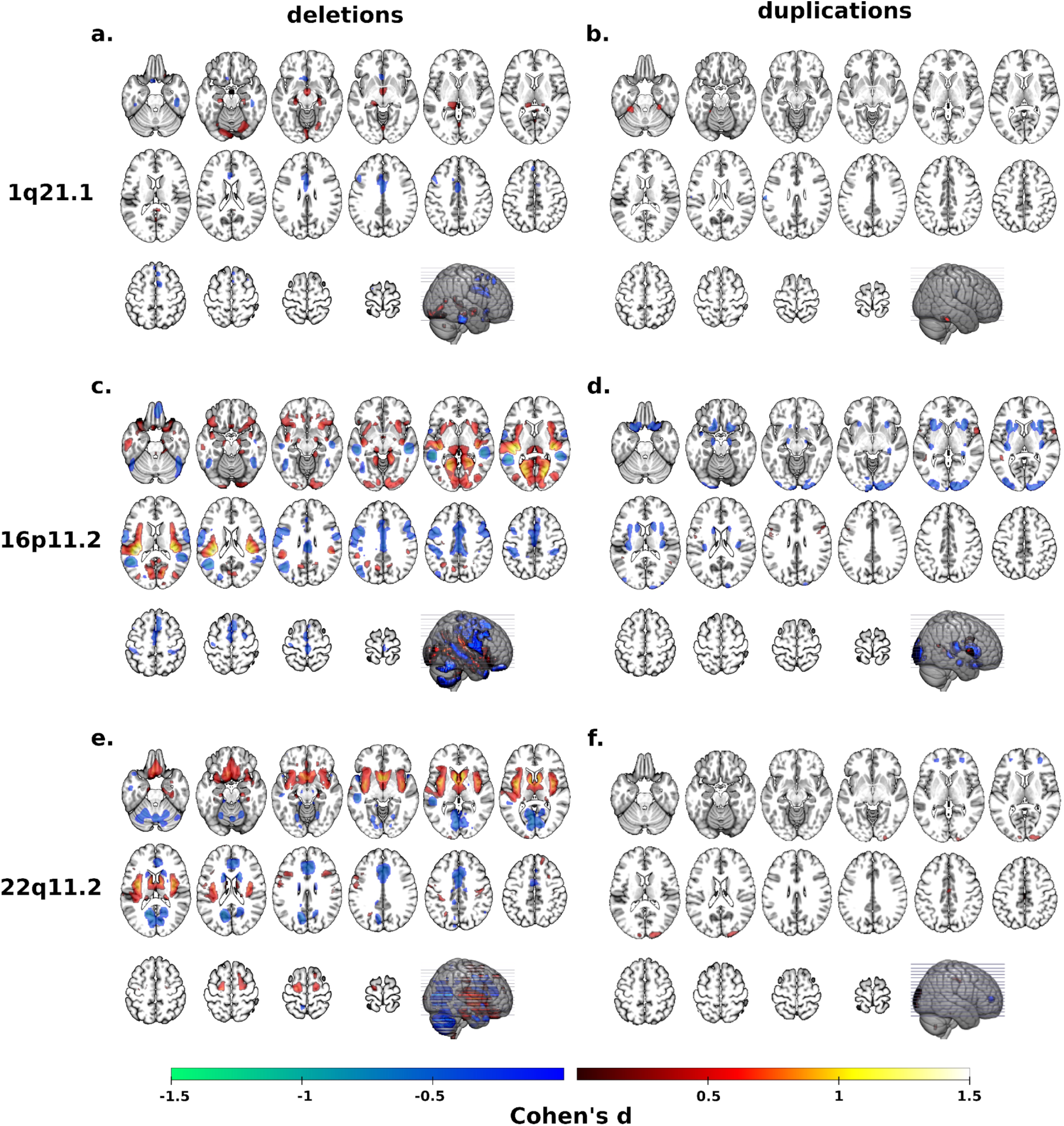
Cohen’s d maps of VBM regional brain differences in deletion and duplication carriers at the 1q21.1, 16p11.2 and 22q11.2 loci compared to controls. Regional brain differences adjusted for total grey matter volume. Left and right columns show results for deletions (a, c, e) and duplication (b, d, f) carriers respectively. Color maps show the significant effects of each CNV, thresholded at q< 0.05 FWE. Color scale represents positive and negative Cohen’s d effect sizes were estimated. Linear model were adjusted for sex, linear and quadratic expansion of age and total grey matter volume. 15q11.2 was not displayed because only a few voxels survived FWE correction.

The conjunction analysis of 3 deletion *vs* control contrasts showed overlap in the left and right middle cingulate gyrus (Figure 4a), corresponding to the 8^th^ and 18^th^ highest loading regions on the first CCA dimension. However, conjunction analyses of FWE thresholded maps are extremely stringent and constrained by the group with the smallest effect and sample size. For example, in the 1q21.1 deletion group we could identify 1.5% of grey matter voxels that survived FWE correction, while the large subject sample and effect-size of the 22q11.2 deletions allowed for the identification of alterations in 14.8% of grey matter (supplementary eTable 2).

**Figure 4:**
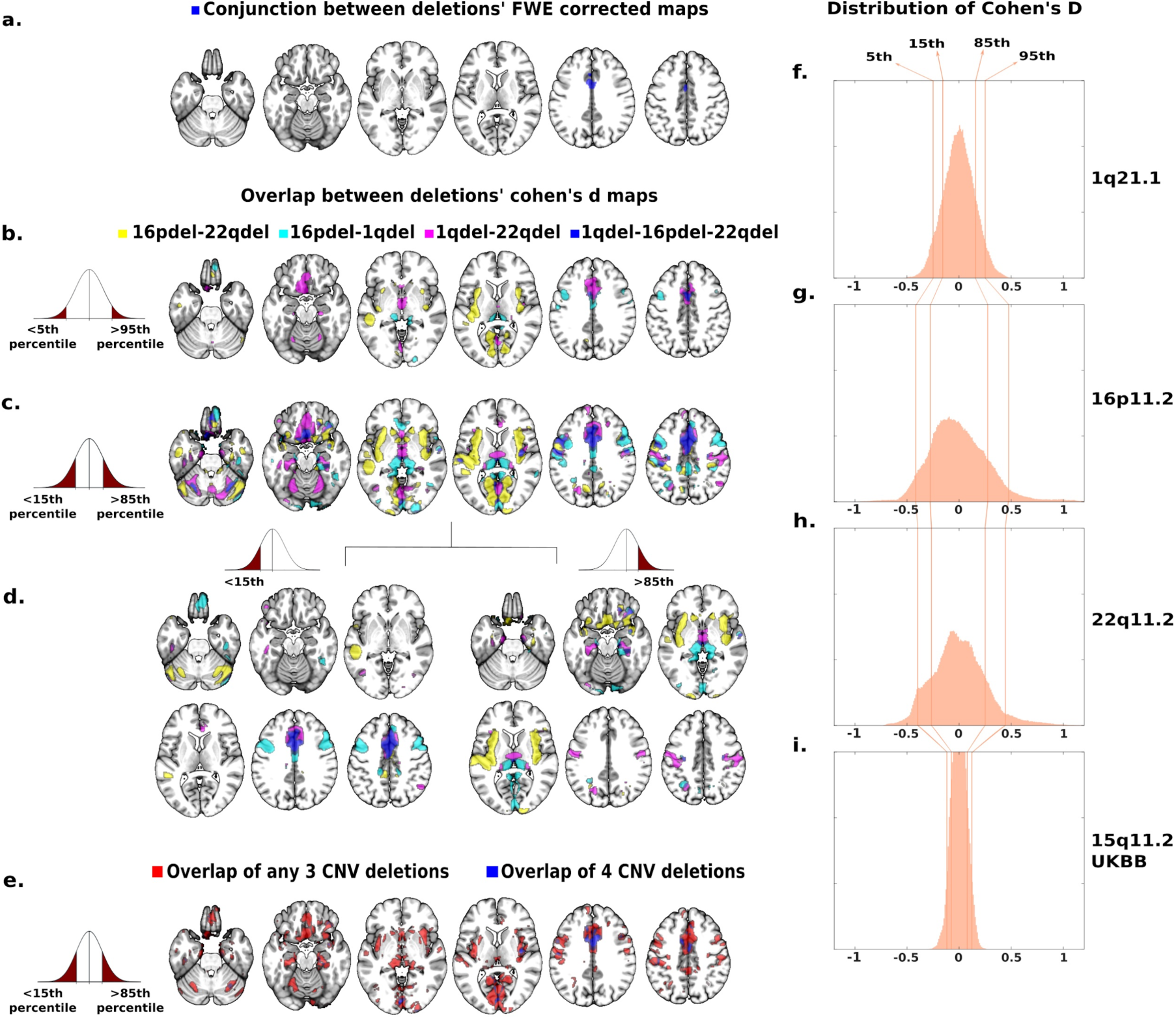
Spatial overlap across deletions at 3 and 4 genomic loci. (a-d) The spatial overlap between the grey matter volume effects of 3 clinically ascertained deletions (16p11.2, 1q21.1 and 22q11.2). (a) The blue map is the conjunction of 3 FWE corrected (q< 0.05) contrasts between deletion and controls, demonstrating that volume in the middle cingulate gyrus is significantly decreased in all 3 deletions. (b-d) Spatial overlap between 3 clinically ascertained deletions adjusted for effect size and sample size by using thresholds based on ranking: <5^th^/>95^th^ (b) and <15^th^/>85^th^ (c) percentiles of Cohen’s d values. Spatial overlap is shown separately for the top negative and positive Cohen’s d values (d). Overlap across the 3 clinically ascertained deletions are reported in blue and pairwise overlaps are reported in purple, cyan and yellow. (e) Spatial overlap across clinically and non-clinically ascertained deletions at 4 genomic loci for <15^th^ and >85^th^ percentile of Cohen’s d values. Overlap of all four deletions is shown in blue. Overlaps of any combination of three deletions are shown in red. Top ranking Cohen’s d values used in (b, c, d, e) are presented on the density plots for all four deletions: 1q21.1 (f), 16p11.2 (g), 22q11.2 (h) and 15q11.2 (i). The X axes values of the 3 density plots are Cohen’s d. Corresponding maps for duplications as well as Surface Area and Cortical Thickness are reported in supplemental eFigures 5-10.

Therefore, to identify shared patterns across CNVs we ranked Cohen’s d maps and overlapped voxels with similar rankings. The conservative group of voxels with Cohen’s d values <5^th^ and >95^th^ percentiles, showed spatial overlap across all deletions in the middle cingulate gyrus (p-valueSHUFFLE<10e-4, Figure 4b). Using a lenient threshold for voxels with Cohen’s d <15^th^ and >85^th^ percentiles, we observed a broader overlap between deletions (p-valueSHUFFLE<10e-4, Figure 4c). Volumes of the middle and anterior cingulate extending to the medial frontal cortex and supplementary motor cortex were decreased in all deletions while volumes were increased in the thalamus, ventral diencephalon, orbital gyrus and parahippocampal gyrus (Figure 4d). Supplementary motor cortex, posterior insula, orbital gyrus, precentral gyrus and thalamus were also found in the top 20 contributors of the CCA dimensions 1. Cerebellum and anterior insula were found in the top 20 contributors of CCA dimension 2 (Figure 2b, supplementary eTable 5).

Spatial convergence could be related to clinical ascertainment. We, therefore, recomputed the deletion convergence map by replacing the clinically ascertained 1q21.1 Cohen’s d map by the one calculated using 1q21.1 deletion carriers from the UK Biobank (Table 1). This new deletion convergence map was similar to the initial one presented above with a dice index of 39.4% (p-valueSPIN< 10e-4).

Finally, we investigated the robustness of the deletion convergence map by intersecting it with a fourth map calculated with 72 carriers of 15q11.2 deletions and 965 controls from the UK Biobank (supplementary eTable 2, Figure 4i). Voxels with Cohen’s d values <15^th^ and >85^th^ percentiles significantly overlapped with the clinically ascertained deletion convergence pattern (p-valueSPIN <10e-4, Figure 4e). We did not perform these analyses for 16p11.2 and 22q11.2 due to the limited sample size in UKBB.

Comparable findings were observed in the analysis, performed in parallel, for Freesurfer derived SA and CT measures (supplementary eFigure 3-4; 6-10).

### 4. Duplications at 4 genomic loci converge on common neuroanatomical alterations

Conjunction analysis of the FWE thresholded duplication maps at the 1q21.1, 16p11.2 and 22q11.2 loci showed no overlap (Figure 3d,f), but we were underpowered to detect the individual effects of 1q21.1 and 22q11.2 duplications (supplementary eTable 2). We therefore tested spatial overlap of voxels with Cohen’s d values <15^th^ and >85^th^ percentiles. Spatial overlap across all three clinically ascertained duplications (p-valueSHUFFLE<10e-4, supplementary eFigure 5b) was characterized by larger volume in the middle cingulate gyrus, fusiform gyrus, superior temporal gyrus and supplementary motor cortex and smaller volume in anterior insula, caudate and temporal pole. These regions were among the top 20 contributors of both CCA dimensions. (Figure 2b, supplementary eTable 5).

We carried out the same sensitivity analyses performed above for deletions. The new duplication convergence map including 1q21.1 duplications and controls from the UKBB showed significant overlap with the initial duplication convergence map (dice-index of 26%, p-valueSPIN <10e-4). The intersection between the duplication convergence map above and a fourth map computed with 76 15q11.2 duplication carriers and 965 controls from the UK Biobank (supplementary eTable 2, eFigure 5h) also showed significant overlap (p-valueSPIN <10e-4, eFigure 5d).

Comparable findings were observed for Freesurfer derived SA and CT measures (supplementary eFigure 3-4; 7-10).

### 5. Effect size ratio and mirror effects of deletions and duplications

Deletions had larger effect sizes on grey matter alterations compared to duplications. Cohen’s d distributions showed a 1.24 to 2-fold deletion/duplication ratio, F test, p<10e-16 (Figure 1, supplementary eTable 11). Similar effect-size ratio is also observed for SA alterations (supplementary eTable9). This was not the case for 15q11.2 deletions and duplications, which showed equally small effect sizes.

We investigated the contrasting regional effects of deletion and duplication on brain morphometry. Deletion and duplication carriers showed brain-wide regional mirror effects for regional volumes and SA, but not CT, at all 4 genomic loci (Figure 5, supplementary eTable 8-10). The strongest anticorrelation of Cohen’s d values was observed for 16p11.2 (p-valueSPIN<10e-4) followed by 15q11.2 (p-valueSPIN<10e-4), 1q21.1 (p-valueSPIN<0.033) and 22q11.2 (p-valueSPIN<0.038). This was true in clinically and non-clinically ascertained CNV carriers (supplementary eTable 6-8).

**Figure 5:**
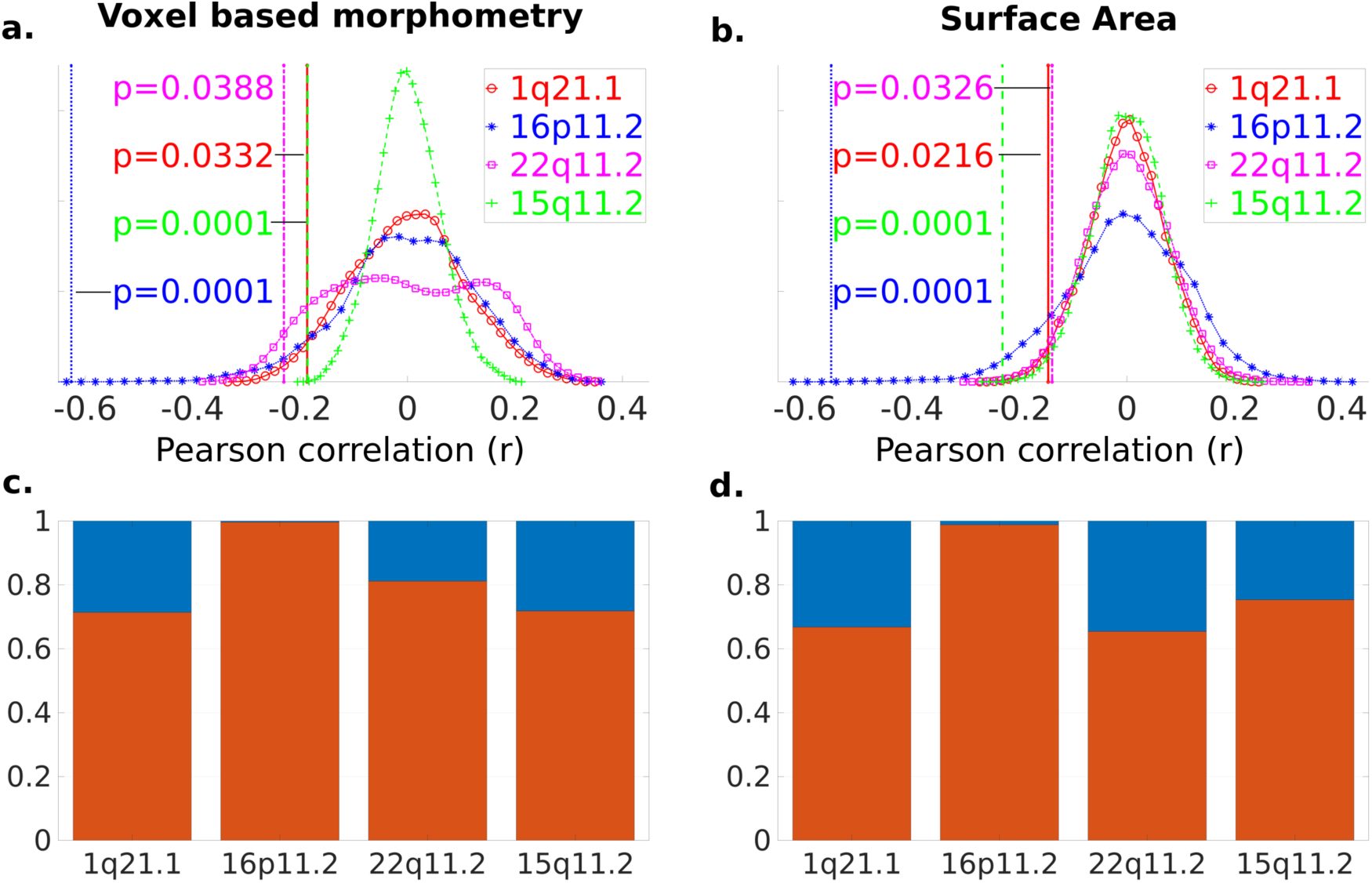
Cortex-wide mirror effects between deletions and duplications. Pearson correlations between Cohen’s d values for deletions and duplications. Voxel-Based Morphometry (a) and Surface Area (b) are adjusted for total gray matter and total surface area respectively. The 4 vertical lines represent the correlation (Pearson r) between deletions and duplications at each locus: 1q21.2 (red), 15q11.2 (green), 16p11.2 (blue), and 22q11.2 (magenta), with the corresponding empirical p-values (uncorrected) shown next to them in same color code. The 4 density plots represent the distribution of Pearson Correlations obtained by performing 10000 spin permutations of duplication maps while keeping deletion maps fixed. Negative correlations between deletions and duplication are observed across loci and are significantly different (Bonferroni) from the null distributions for 16p11.2 and 15q11.2 (p-values are uncorrected). X axis = Pearson r coefficients, y axis = the surface under the curve is 100% of the distribution. (c-d) Mirror effects between deletions and duplications at both tails of the distribution c) The red bar is the proportion of voxels that are in the top 85th percentile for deletions and the lower 15th percentile for duplications and vice versa. The blue bar represents voxels that are either in the top 85th or lower 15th percentile for both deletions and duplications. d) The same bar plots are presented for surface area. (All Correlation values are reported in supplemental eTable 8-10).

### 6. Relationships with previously defined cross-psychiatric disorder maps

We investigated similarities between our CNV convergence maps and alterations reported by a cross-disorder meta-analysis of case-control studies of schizophrenia, bipolar disorder, obsessive-compulsive disorder, substance-use disorder, major depression and anxiety ^39^. Deletion convergence map showed alterations affecting the same regions (cingulate and insula) with contiguous, but only marginally overlapping clusters (supplementary eFigure 11). The duplication convergence map also showed marginal overlaps with a contiguous cluster in the anterior insula.

## Discussion

Our study demonstrates that deletions and duplications at 4 distinct genomic loci have independent effects on global and regional brain morphometry. The Eight CNVs, associated with ASD and SZ, affect regional brain volumes along two gene-morphometry dimensions that emerged from our CCA results. These dimensions included regions such as the temporal gyrus, calcarine cortex, supplementary motor cortex, insula, middle cingulate gyrus, accumbens, subcallosal area, cuneus, precuneus, brain stem and superior frontal gyrus. Deletions and duplications of the same genomic loci lie on opposite ends of these brain dimensions. Univariate analyses similarly point towards a pattern of differences involving anterior cingulate, posterior insula and supplementary motor cortex observed in deletions across 4 loci. For duplications a different pattern is observed involving middle cingulate, anterior insula and lingual gyrus. Mirror effects are observed at the global and regional levels across loci, for volume and SA but not CT.

### Dissociation between global and regional effects

Systematic comparison across loci suggests that CNVs have independent effect sizes on global and regional brain morphometry. For example, the 1q21.1 deletions and duplications highlight the contrast between very large effects on global measures, with small regional effects adjusted for total GM. The same dissociation is observed between the directionalities of global and regional effects across the 4 genomic loci. As an example, 1q21.1 and 22q11.2 deletions have negative effects on TIV and GM, while 16p11.2 deletions have positive effects, and 15q11.2 deletions have no detectable effects. In contrast, all four deletions show smaller regional volumes in the middle and anterior cingulate, medial frontal cortex and supplementary motor cortex, as well as larger volumes in the thalamus, orbital gyrus and parahippocampal gyrus. The same dissociation between global and regional effect sizes and between directionalities applies to duplications.

We posit that global and local effects may be mechanistically unrelated. Animal studies have proposed that CNV-related differences in global brain volume may be due to the modulation of embryonic neurogenesis ^6^, dendrite growth or spine and synapse development ^23^. Larger brain size in individuals diagnosed with autism-related genetic conditions such as tuberous sclerosis complex has been linked to cell body size and is sensitive to medication ^40^. On the other hand, the mechanisms underlying local relative effects have not been investigated in animal models. Although both, global and regional brain alterations show mirror effects in deletions and duplications across most loci.

### General effects of gene dosage on brain structure

Altering gene dosage at 4 distinct loci encompassing 60, 29, 12 and 4 genes lead to a degree of shared regional patterns. In line with this observation, a complex landscape of rare deleterious CNVs have been associated with cognition ^21^, increased risk of neurodevelopmental and psychiatric disorders such as autism ^1,20,22^ and schizophrenia ^2^. Recent studies estimated that 71%–100% of any 1-MB window in the human genome contributes to increased risk for schizophrenia ^41^. The same has been demonstrated for autism ^22^. We therefore speculate that CNVs will lead to patterns of brain alterations similar to the one characterized in our study irrespective of their genomic location.

A plausible hypothesis for the pervasive effects of CNVs on cognition, behavior and brain architecture, is that these phenomena are related to emerging properties of the genome, rather than the function of individual genes ^42^. In other words, gene dosage at any node of the genomic network will alter its efficiency leading to a measurable effect on brain organization and behavior. Gene dosage affecting diverse molecular functions may lead to a limited number of ways in which the brain reconfigures, compared to non-carriers.

### Neuroanatomical patterns across brain measures

Systematic analysis through the two most widespread computational neuroanatomy frameworks (VBM and Freesurfer) shows that main grey matter volume findings are recapitulated by SA results. On the other hand, changes in CT seem to be distinct, which is in line with previous investigations of 16p11.2 CNVs ^15^ as well as studies suggesting that apparent CT is mainly related to cortical myelination ^43^. Volume is a product of SA and CT, measures that have been shown to be genetically unrelated ^44^.

### Limitations

Our study was focused on investigating shared features across genomic loci. The study of many more loci would be required to characterize specific effects. Integrating data from multiple study sites may have introduced noise and heterogeneity in our investigation. We have previously shown that, although site effects on neuroimaging data are measurable, they do not influence the neuroanatomical patterns associated with CNVs at the 16p11.2, 22q11.2 and 15q11.2 loci ^10,15,45^.

We carried out the first whole-brain voxel- and vertex-wise analyses for 15q11.2 and 1q21.1 CNVs and replicated findings for 22q11.2 and 16p11.2 CNVs ^10,14,17^. For 1q21.1, we used multisite data as an opportunity to perform sensitivity analyses and challenge the robustness of our results. Although we were underpowered to clearly delineate the small effect of 15q11.2 deletions and duplications at the voxel level, regions with top Cohen’s d values that significantly overlap with the 3 other loci –frontal, cingulate gyrus, and parietal lobes–, are concordant with those reported in a recent large scale study using the Desikan-Killiany Atlas ^45^.

### Conclusions

In this proof of concept, our investigation demonstrates the relevance of simultaneously analysing the effect of several genomic variants on neuroimaging intermediate phenotypes. Extending our approach to the rapidly expanding number of rare genomic variants associated with psychiatric disorders should provide the field with a scheme to understand general principles linking deleterious genomic variants with their consequence on human brain architecture. These general neuroanatomical patterns may help understand the broad locus heterogeneity of psychiatric conditions.

## Data Availability

Overlap maps are available through a request to the corresponding authors.

## Founding

This research was supported by Calcul Quebec (http://www.calculquebec.ca) and Compute Canada (http://www.computecanada.ca), the Brain Canada Multi-Investigator initiative, the Canadian Institutes of Health Research, CIHR_400528, The Institute of Data Valorization (IVADO) through the Canada First Research Excellence Fund, Healthy Brains for Healthy Lives through the Canada First Research Excellence Fund. Dr Jacquemont is a recipient of a Canada Research Chair in neurodevelopmental disorders, and a chair from the Jeanne et Jean Louis Levesque Foundation. The Cardiff CNV cohort was supported by the Wellcome Trust Strategic Award “DEFINE” and the National Centre for Mental Health with funds from Health and Care Research Wales (code 100202/Z/12/Z). The CHUV cohort was supported by the SNF (Maillard Anne, Project, PMPDP3 171331). Data from the UCLA cohort provided by Dr. Bearden (participants with 22q11.2 deletions or duplications and controls) was supported through grants from the NIH (U54EB020403), NIMH (R01MH085953, R01MH100900, R03MH105808), and the Simons Foundation (SFARI Explorer Award). Claudia Modenato was supported by the doc.mobility grant provided by the Swiss National Science Foundation (SNSF). Kuldeep Kumar was supported by The Institute of Data Valorization (IVADO) Postdoctoral Fellowship program, through the Canada First Research Excellence Fund. Dr. Bzdok was supported by the Healthy Brains Healthy Lives initiative (Canada First Research Excellence fund), and by the CIFAR Artificial Intelligence Chairs program (Canada Institute for Advanced Research).

## Author contributions

C.Mod., K.K., B.D. and S.J. designed the study, analysed imaging data and drafted the manuscript.

### Analyses

C.Mod. and K.K. did all the preprocessing and analysis of neuroimaging data, DB provided scripts and mentored the CCA analysis. C.Mor., C.E.B. and D.B. contributed in result interpretation and in the editing of the manuscript.

### Data collection

C.Mod., A.M., A.P., S.R. and S.M-B. recruited and scanned participants in the 16p11.2 European Consortium. S.L., C.O.M., N.Y., P.T., E.D., F. T-D., V.C., A.R.C., F.D. recruited and scanned participants in the Brain Canada cohort. L.K. collected and provided the data for the UCLA cohort. D.E.J.L., M.J.O., M.B.M. V.d.B., J.H. and A.I.S., provided the data for the Cardiff cohort.

All authors provided feedback on the manuscript.

## Article Informations

Contributors to the Simons VIP Consortium include the following: Hanalore Alupay, BS, Benjamin Aaronson, BS, Sean Ackerman, MD, Katy Ankenman, MSW, Ayesha Anwar, BA, Constance Atwell, PhD, Alexandra Bowe, BA, Arthur L. Beaudet, MD, Marta Benedetti, PhD, Jessica Berg, MS, Jeffrey Berman, PhD, Leandra N. Berry, PhD, Audrey L. Bibb, MS, Lisa Blaskey, PhD, Jonathan Brennan, PhD, Christie M. Brewton, BS, Randy Buckner, PhD, Polina Bukshpun, BA, Jordan Burko, BA, Phil Cali, EdS, Bettina Cerban, BA, Yishin Chang, MS, Maxwell Cheong, BE, MS, Vivian Chow, BA, Zili Chu, PhD, Darina Chudnovskaya, BS, Lauren Cornew, PhD, Corby Dale, PhD, John Dell, BS, Allison G. Dempsey, PhD, Trent Deschamps, BS, Rachel Earl, BA, James Edgar, PhD, Jenna Elgin, BS, Jennifer Endre Olson, PsyD, Yolanda L Evans, MA, Anne Findlay, MA, Gerald D Fischbach, MD, Charlie Fisk, BS, Brieana Fregeau, BA, Bill Gaetz, PhD, Leah Gaetz, MSW, BSW, BA, Silvia Garza, BA, Jennifer Gerdts, PhD, Orit Glenn, MD, Sarah E Gobuty, MS, CGC, Rachel Golembski, BS, Marion Greenup, MPH, MEd, Kory Heiken, BA, Katherine Hines, BA, Leighton Hinkley, PhD, Frank I. Jackson, BS, Julian Jenkins III, PhD, Rita J. Jeremy, PhD, Kelly Johnson, PhD, Stephen M. Kanne, PhD, Sudha Kessler, MD, Sarah Y. Khan, BA, Matthew Ku, BS, Emily Kuschner, PhD, Anna L. Laakman, MEd, Peter Lam, BS, Morgan W. Lasala, BA, Hana Lee, MPH, Kevin LaGuerre, MS, Susan Levy, MD, Alyss Lian Cavanagh, MA, Ashlie V. Llorens, BS, Katherine Loftus Campe, MEd, Tracy L. Luks, PhD, Elysa J. Marco, MD, Stephen Martin, BS, Alastair J. Martin, PhD, Gabriela Marzano, HS, Christina Masson, BFA, Kathleen E. McGovern, BS, Rebecca McNally Keehn, PhD, David T. Miller, MD, PhD, Fiona K. Miller, PhD, Timothy J. Moss, MD, PhD, Rebecca Murray, BA, Srikantan S. Nagarajan, PhD, Kerri P. Nowell, MA, Julia Owen, PhD, Andrea M. Paal, MS, Alan Packer, PhD, Patricia Z. Page, MS, Brianna M. Paul, PhD, Alana Peters, BS, Danica Peterson, MPH, Annapurna Poduri, PhD, Nicholas J. Pojman, BS, Ken Porche, MS, Monica B. Proud, MD, Saba Qasmieh, BA, Melissa B. Ramocki, MD, PhD, Beau Reilly, PhD, Timothy P. L. Roberts, PhD, Dennis Shaw, MD, Tuhin Sinha, PhD, Bethanny Smith-Packard, MS, CGC, Anne Snow Gallagher, PhD, Vivek Swarnakar, PhD, Tony Thieu, BA, MS, Christina Triantafallou, PhD, Roger Vaughan, PhD, Mari Wakahiro, MSW, Arianne Wallace, PhD, Tracey Ward, BS, Julia Wenegrat, MA, and Anne Wolken, BS. Members of the European 16p11.2 Consortium include the following: Addor Marie-Claude, Service de génétique médicale, Centre Hospitalier Universitaire Vaudois, Lausanne University, Switzerland; Alexandre Raymond, Center for Integrative Genomics, Lausanne University, Switzerland; Andrieux Joris, Institut de Génétique Médicale, CHRU de Lille, Hopital Jeanne de Flandre, France; Arveiler Benoît, Service de génétique médicale, CHU de Bordeaux-GH Pellegrin, France; Baujat Geneviève, Service de Génétique Médicale, CHU Paris - Hôpital Necker-Enfants Malades, France; Sloan-Béna Frédérique, Service de médecine génétique, Hôpitaux Universitaires de Genève - HUG, Switzerland; Belfiore Marco, Service de génétique médicale, Centre Hospitalier Universitaire Vaudois, Lausanne University, Switzerland; Bonneau Dominique, Service de génétique médicale, CHU d’Angers, France; Bouquillon Sonia, Institut de Génétique Médicale, Hopital Jeanne de Flandre, Lille, France; Boute Odile, Hôpital Jeanne de Flandre, CHRU de Lille, Lille, France; Brusco Alfredo, Genetica Medica, Dipartimento di Scienze Mediche, Università di Torino, Italy; Busa Tiffany, Département de génétique médicale, CHU de Marseille, Hôpital de la Timone, France; Caberg Jean-Hubert, Centre de génétique humaine, CHU de Liège, Belgique; Campion Dominique, Service de psychiatrie, Centre hospitalier de Rouvray, Sotteville lès Rouen, France; Colombert Vanessa, Service de génétique médicale, Centre Hospitalier Bretagne Atlantique CH Chubert-Vannes, France; Cordier Marie-Pierre, Service de génétique clinique, CHU de Lyon, Hospices Civils de Lyon, France; David Albert, Service de Génétique Médicale, CHU de Nantes, Hôtel Dieu, France; Debray François-Guillaume, Service de Génétique Humaine, CHU Sart Tilman - Liège, Belgique; Delrue Marie-Ange, Service de génétique médicale, CHU de Bordeaux, Hôpital Pellegrin, France; Doco-Fenzy Martine, Service de Génétique et Biologie de la Reproduction, CHU de Reims, Hôpital Maison Blanche, France; Dunkhase-Heinl Ulrike, Department of Pediatrics, Aabenraa Hospital, Sonderjylland, Denmark; Edery Patrick, Service de génétique clinique, CHU de Lyon, Hospices Civils de Lyon, France; Fagerberg Christina, Department of Clinical Genetics, Odense University hospital, Denmark; Faivre Laurence, Centre de génétique, Hôpital d’Enfants, CHU Dijon Bourgogne - Hôpital François Mitterrand, France; Forzano Francesca, Ambulatorio di Genetica Medica, Ospedali Galliera di Genova, Italy and Clinical Genetics Department, 7^th^ Floor Borough Wing, Guy’s Hospital, Guy’s & St Thomas’ NHS Foundation Trust, Great Maze Pond, London SE1 9RT, UK; Genevieve David, Département de Génétique Médicale, Maladies Rares et Médecine Personnalisée, service de génétique clinique, Université Montpellier, Unité Inserm U1183, CHU Montpellier, Montpellier, France; Gérard Marion, Service de Génétique, CHU de Caen, Hôpital Clémenceau, France; Giachino Daniela, Genetica Medica, Dipartimento di Scienze Cliniche e Biologiche, Università di Torino, Italy; Guichet Agnès, Service de génétique, CHU d’Angers, France; Guillin Olivier, Service de psychiatrie, Centre hospitalier du Rouvray, Sotteville lès Rouen, France; Héron Delphine, Service de Génétique clinique, CHU Paris-GH La Pitié Salpêtrière-Charles Foix - Hôpital Pitié Salpêtrière, France; Isidor Bertrand, Service de Génétique Médicale, CHU de Nantes, Hôtel Dieu, France; Jacquette Aurélia, Service de Génétique clinique, CHU Paris-GH La Pitié Salpêtrière-Charles Foix - Hôpital Pitié-Salpêtrière, France; Jaillard Sylvie, Service de Génétique Moléculaire et Génomique – Pôle biologie, CHU de Rennes, Hôpital Pontchaillou, France; Journel Hubert, Service de génétique médicale, Centre Hospitalier Bretagne Atlantique CH Chubert-Vannes, France; Keren Boris, Centre de Génétique Moléculaire et Chromosomique, CHU Paris-GH La Pitié Salpêtrière-Charles Foix - Hôpital Pitié-Salpêtrière, France; Lacombe Didier, Service de génétique médicale, CHU de Bordeaux-GH Pellegrin, France; Lebon Sébastien, Pediatric Neurology Unit, Department of Pediatrics, Lausanne University Hospital, Lausanne, Switzerland; Le Caignec Cédric, Service de Génétique Médicale - Institut de Biologie, CHU de Nantes, France; Lemaître Marie-Pierre, Service de Neuropédiatrie, Centre Hospitalier Régional Universitaire de Lille, France; Lespinasse James, Service génétique médicale et oncogénétique, Hotel Dieu, Chambéry, France; Mathieu-Dramart Michèle, Service de Génétique Clinique, CHU Amiens Picardie, France; Mercier Sandra, Service de Génétique Médicale, CHU de Nantes, Hôtel Dieu, France; Mignot Cyril, Service de Génétique clinique, CHU Paris-GH La Pitié Salpêtrière-Charles Foix - Hôpital Pitié-Salpêtrière, France; Missirian Chantal, Département de génétique médicale, CHU de Marseille, Hôpital de la Timone, France; Petit Florence, Service de génétique clinique Guy Fontaine, Hôpital Jeanne de Flandre, CHRU de Lille, France; Pilekær Sørensen Kristina, Department of Clinical Genetics, Odense University Hospital, Denmark; Pinson Lucile, Département de Génétique Médicale, Maladies Rares et Médecine Personnalisée, service de génétique clinique, Université Montpellier, Unité Inserm U1183, CHU Montpellier, Montpellier, France; Plessis Ghislaine, Service de Génétique, CHU de Caen, Hôpital Clémenceau, France; Prieur Fabienne, Service de génétique clinique, CHU de Saint-Etienne - Hôpital Nord, France; Rooryck-Thambo Caroline, Laboratoire de génétique moléculaire, CHU de Bordeaux-GH Pellegrin, France; Rossi Massimiliano, Service de génétique clinique, CHU de Lyon, Hospices Civils de Lyon, France; Sanlaville Damien, Laboratoire de Cytogénétique Constitutionnelle, CHU de Lyon, Hospices Civils de Lyon, France; Schlott Kristiansen Britta, Department of Clinical Genetics, Odense University Hospital, Denmark; Schluth-Bolard Caroline, Laboratoire de Cytogénétique Constitutionnelle, CHU de Lyon, Hospices Civils de Lyon, France; Till Marianne, Service de génétique clinique, CHU de Lyon, Hospices Civils de Lyon, France; Van Haelst Mieke, Department of Genetics, University Medical Center Utrecht, Holland; Van Maldergem Lionel, Centre de Génétique humaine, CHRU de Besançon - Hôpital Saint-Jacques, France.

